# Clinical Decision Support Using a Partially Instantiated Probabilistic Graphical Model Optimized through Quantum Annealing: Proof-of-Concept of a Computational Method Using a Clinical Data Set

**DOI:** 10.1101/2020.12.13.20248138

**Authors:** David K. Sahner, Richard J. Williams

## Abstract

An approach is described to building a clinical decision support tool which leverages a partially instantiated graphical model optimized through quantum annealing. Potential advantages of such a strategy include the practical, and potentially real-time, use of multidimensional patient data to make a host of intuitively understandable predictions and recommendations in complex cases which are informed by a data-driven probabilistic model. Preliminary proof-of-concept of the general approach is demonstrated using a large well-established anonymized patient data set, revealing the predictive capability of a specific model. Ideas for future research are discussed.

## I. Introduction

Although astonishing scientific and medical advances have transformed both our understanding of human physiology and the potential to help patients, the complexity of medicine and biology has also begun to outstrip the capacity of any single human intellect. Primary care physicians, overburdened and awash in data and exploding medical information, are faced daily with the daunting challenge of caring for complex patients with multiple conditions. Machine learning techniques and applications may hold promise in various health care-related settings^1^, but current predictive medical decision support tools for diagnosis, prognosis or risk stratification based on classical machine learning often provide focused predictions for a specific condition, such as sepsis or diabetes mellitus, outcomes in Parkinson’s disease^2^ or response to a statin^3^. What may be more useful is a support system that provides timely wide-ranging insights into a host of risks and likely therapeutic outcomes in specific patients, enabling more accurate diagnosis and intervention, more precise and customized treatment selection, and, by extension, improved patient outcomes in a broad capacity. With the increasing number of aging patients with multiple co-morbidities who are treated with complicated polypharmacy, the progressive increase in medical expenditures, and an increasing focus on quality, such an innovation may be especially helpful in clinical practice.

Probabilistic graphical models have been evaluated as potential clinical decision aids^4,5^, but maximum a posteriori estimation (MAP inference) techniques to “fill in the blanks” in probabilistic models, assigning the most likely value of a clinical variable in a specific case (e.g., diagnosis, therapeutic outcome, etc.) may be hamstrung by the potentially large number of nodes in a model and the NP hard nature of MAP inference. Historically, MAP inference approximation techniques are often slow or scale poorly, prompting interest in novel inference strategies^6^. Although various methods have been employed to render these problems more tractable, large densely connected graphs with many hundreds or thousands of nodes, including large clique sizes, may remain resistant to classical techniques. It is hypothesized that quantum annealing, which is useful in optimization, may offer a solution to this problem. One of the major advantages of this graphical approach, apart from the potential improved feasibility related to quantum efficiencies, is the ability to leverage a large body of existing population-based data, as well as available individual patient-specific data, to make a number of inferences in a given case that are based on a holistic representation of the relationships among numerous variables. In contrast to a black-box deep neural net which is characterized by hidden data representations (abstractions) that may defy human understanding, the outputs of probabilistic graphical models are also explainable. For example, a physician can simply scan the known and inferred values of the nodes in a Markov blanket investing a node whose value has been predicted in order to understand the most proximal sources of relevant information in the model that had direct bearing on that prediction.

This paper envisions a clinical decision support tool based on partially instantiated undirected graphical models in which hundreds or potentially thousands of nodes represent signs, symptoms, diagnoses, laboratory and imaging results, known co-morbidities, demographics, concomitant medications, outcomes and selected genomic, epigenetic, transcriptomic, metabolomic, and proteomic biomarkers, as well as other variables. The weights or potential functions between nodes may be informed by large scale data derived from electronic health records, insurance claims data, clinical studies, and, potentially, other sources. In a given case (i.e., for a single patient), the values of some of the nodes in the graph, essentially a Markov Random Field^7,8^, are known, whereas others (e.g., diagnosis, likelihood of a favorable response to a specific intervention, etc.) are not. The system solves for the most likely values of these unknown nodes in an efficient fashion, given the totality of the data, the graph structure and graph parameters, yielding clinically actionable information in (close to) real-time by exploiting a novel quantum annealing algorithm. In theory, the system could be seamlessly integrated with the electronic health record and accessed through an intuitive user interface with physician and patient portals.

This paper is organized as follows. In section II, the general methods underlying our approach are described, including the manner in which the Markov Random Field is constructed and parameterized, the objective function, and expression of the latter in a form amenable to optimization using the D-Wave quantum annealing machine. This section also reviews basic elements of the D-wave machine and quantum annealing at a high level. Section III provides details on the objectives of the proof-of-concept (POC) study and methods specific to an evaluation, which relied upon data from patients whose ICU courses at Beth Israel Hospital were captured in the MIMIC III database. Section IV furnishes results of the POC study and Section V includes a discussion and conclusions, with ideas for further research.

## II. General Methods

### Overview of Methods

A brief stepwise overview of the methodology is presented first, to provide a framework for a discussion of some of the methodologic details.

1. An undirected graphical model (Markov Random Field) is built with nodes and edges that capture probabilistic relationships among variables in patients. This is accomplished in two substeps:
  a. First, the topology of the model (i.e., nodes and edges) is constructed (Figure 1). Each node corresponds to a variable and edges (links) are drawn between those nodes that, based on existing medical expertise and scientific knowledge, are thought to be related. By related, we mean the likelihood that variable A is true is influenced by the truth value of variable B, or vice-versa. Node selection was also influenced by available clinical data in the MIMIC-III dataset^9^ that was used to parameterize the model.
  b. Next, the model is parameterized and order-reduced. The potential functions (or weights) that serve as the critical parameters of the model are selected using the pseudo-likelihood approach^8^ based on actual clinical data from a training set, reserving a proportion of the data as a test set (i.e., for the clinical proof-of-concept [POC] described in this paper). These potential functions therefore capture empirically determined probabilistic relationships among connected variables in the clinical POC model, which was based on the MIMIC III data set described below. Order reduction^10^ produces acceptable clique sizes for the following (computational) step.
2. After the model is built and parameterized, individual predictions of the values of “unknown” variables can be made for a given case given what is known about the patient. This, too, proceeds in two sub-steps:
  a. Where data are available, nodes in the model are set to a specific truth value given what is known about a given patient (e.g., signs, symptoms, demographics, laboratory and imaging studies, co-morbidities, concomitant medications, etc.).
  b. A quantum annealing-based algorithm, to be described, which relies upon a specific objective function, is then used to infer the truth values of the uninstantiated nodes (i.e., those lacking known truth values) for a specific patient (e.g., likely diagnosis, reasonable likelihood of response to a specific intervention, progression-free survival of greater than 18 months, etc.). See Figure 1 below for part of a mock network.
3. Although not discussed at length in this paper, the output of the algorithm can be translated into ranked clinically actionable insights for a physician’s consideration and discussion with the patient.

**Figure 1:**
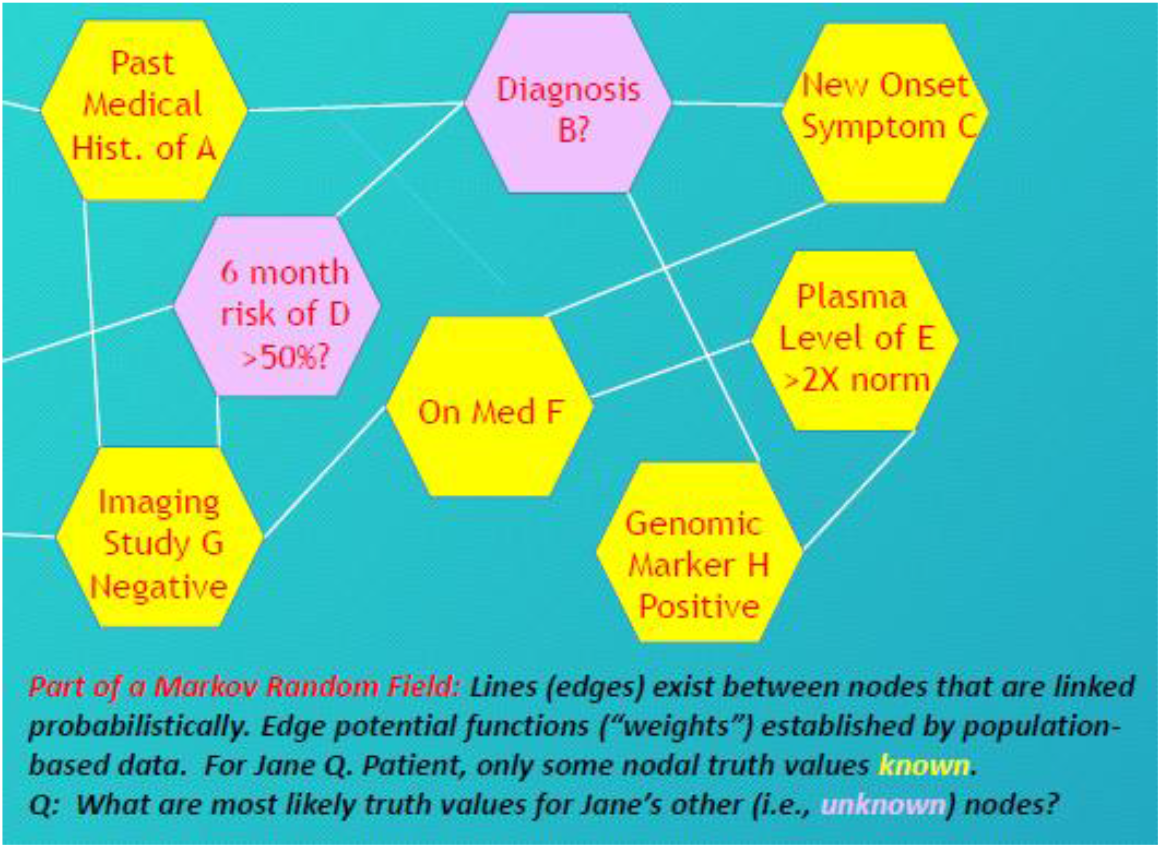
Hypothetical section of the topology of a Markov Random Field used in a model

The use of “Bayesian islands” within a Markov Random Field, an example of which is provided later in Section III, allows the system to classically infer variable values in the context of missing data before the “blanks” in the Markov Random Field are filled in by the quantum annealing algorithm.

### Objective Function for Optimization

The optimization algorithm, which basically infers the values of the “blanks” in the graphic model for a given patient, is driven by quantum annealing. The objective function to be optimized during quantum annealing will first be described, and this will be followed by a brief description of the manner in which quantum annealing optimizes this function. Figure 2 below explains how the objective function is constructed at a high level.

**Figure 2:**
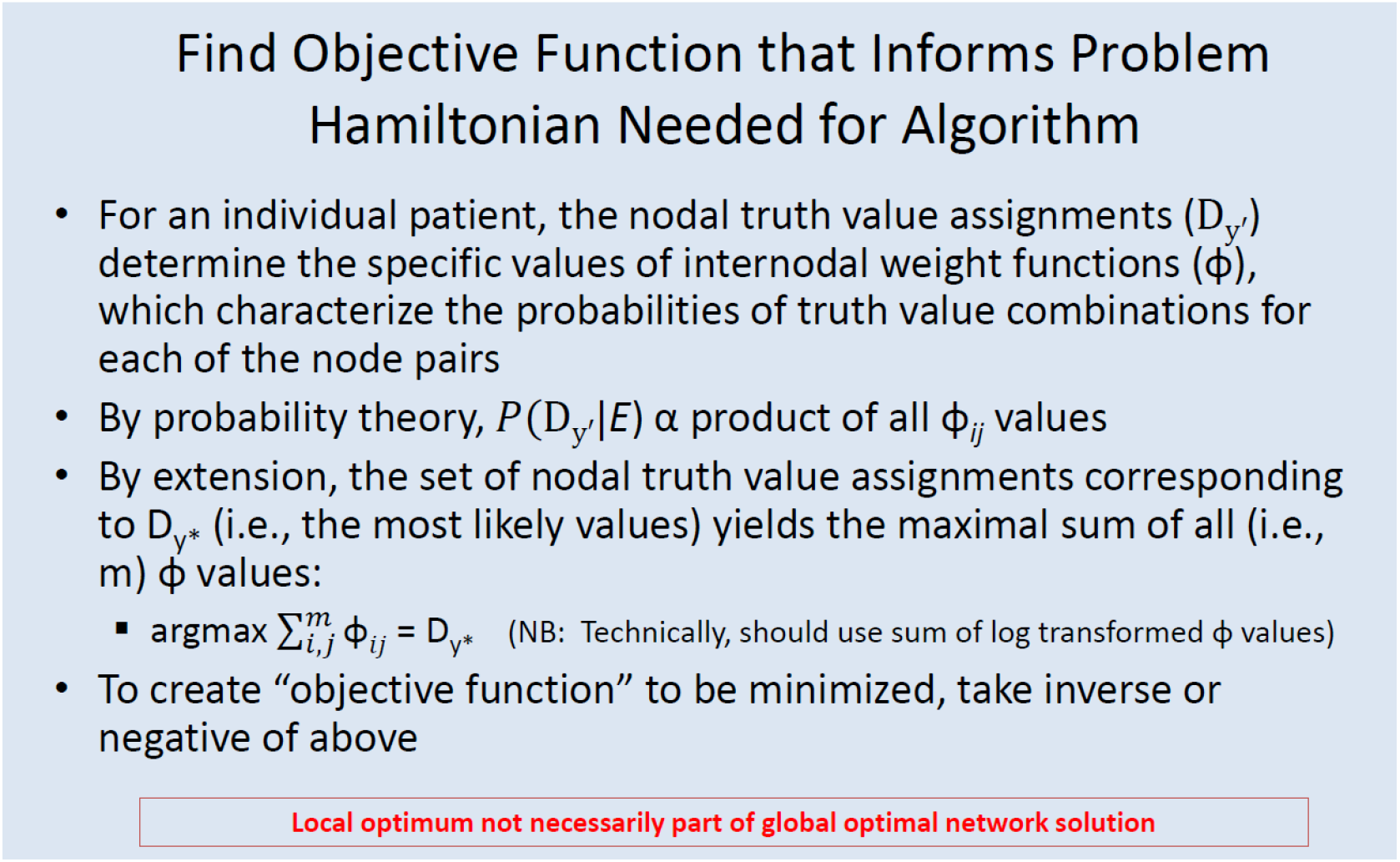
Determining the objective function

Given the connectivity of the D-wave machine, it is necessary to express the potential function (i.e., phi values) between nodes as a function of the truth values of the nodes they join. Each edge in the Markov Random Field is associated with a potential function. This is explained in Figure 3 below.

**Figures 3a and 3b:**
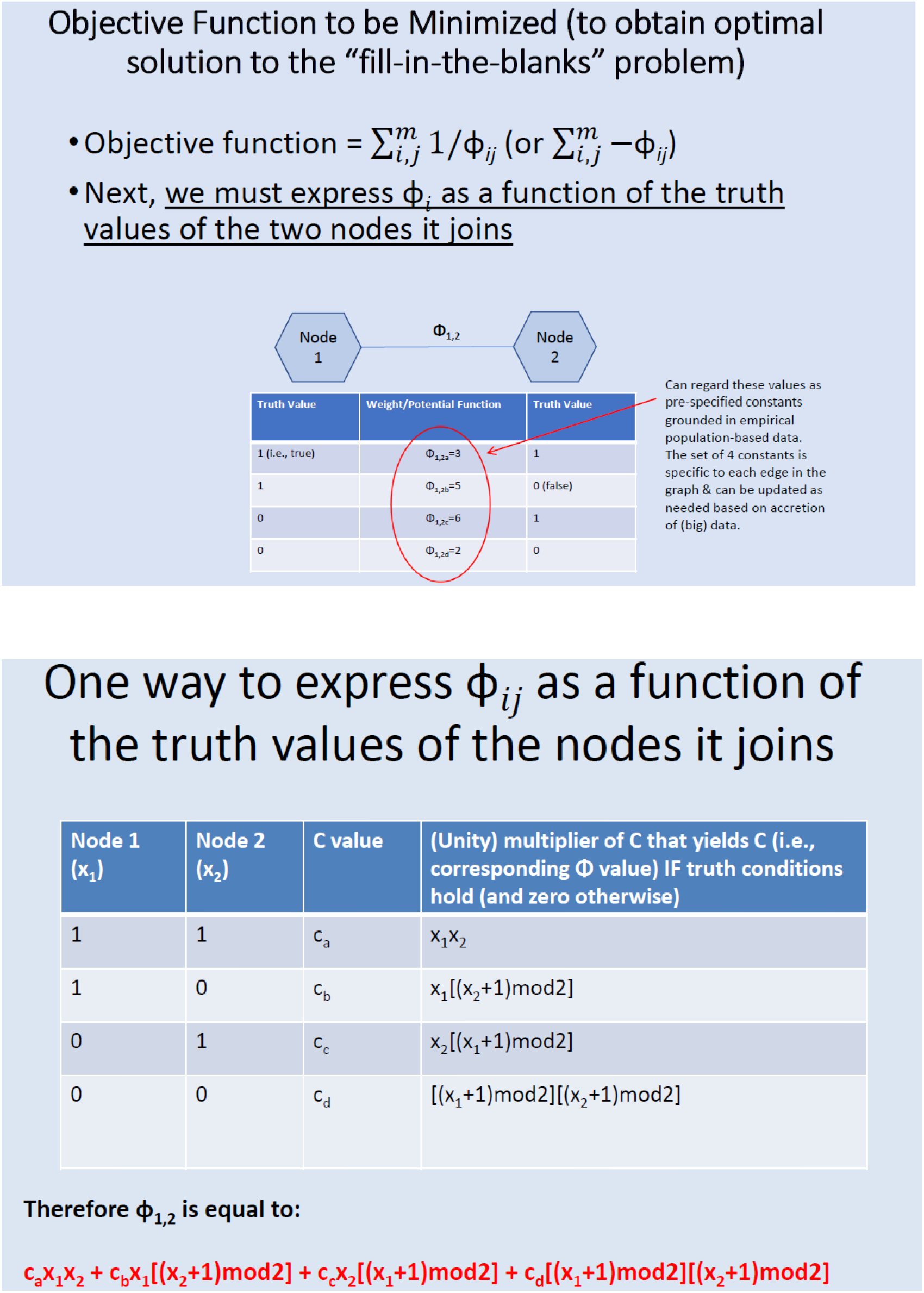
Express phi values between nodes as a function of the truth values of the nodes its corresponding edge joins

If phi values are expressed in this way, then the objective function to be optimized takes the following form:

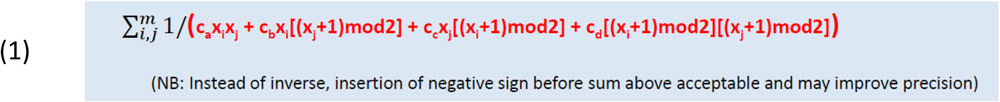

### D-Wave Machine and Adaptation of Objective Function to Quantum Annealing

Quantum annealing^11,12,13,14^ affords a novel approach to solving optimization problems. The D-Wave quantum annealing machine used in our experiments housed up to 2048 functional qubits in a shielded environment. Each qubit consists of a cooled niobium ring that acts as a superconductor which can magnetically encode one of two states or both in superposition. This is analogous to the spin of an electron along the z axis, which, prior to its measurement, may be in a state of quantum superposition (Figure 4). The qubit is not an electron, but mimics the spin of an electron.

**Figure 4:**
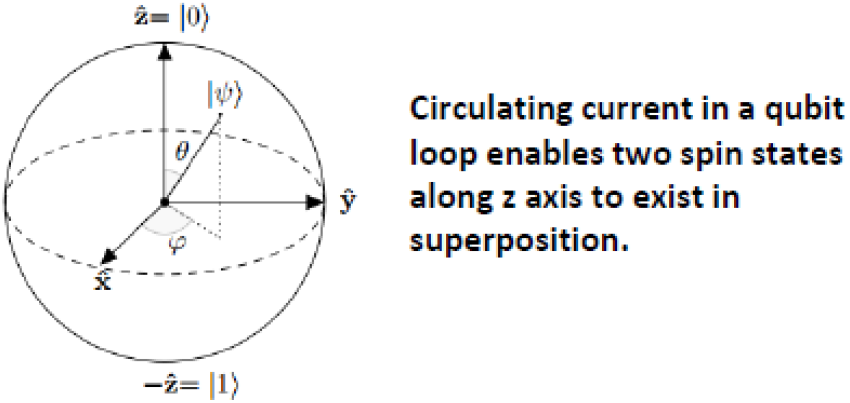
Superposition of spin states

The qubits in the D-wave machine are coupled and arranged in what is known as a chimera architecture. Programmability of the device is enabled by the ability to adjust local magnetic field strengths. To solve a graphical problem of the type outlined in this paper, the graph first must be “embedded” within the architecture of the D-wave machine. If a node in a graphical model has fewer than ∼6 adjacencies (that is, if the order of the node is ∼6 or less), one qubit can be assigned to a single node, but, for nodes with denser connectivity, multiple qubits are required for one logical bit.

At a superficial level, the process of annealing may be thought of as the controlled “ushering” or nudging of an initial Hamiltonian toward a final problem Hamiltonian in or near the ground state. This low energy state encodes the solution to the inference problem in the spins of the qubits. In essence, the measured eigenvalues of those spins (either 0 or 1), obtained after the annealing run is complete, are used to compute the likely truth values of the nodes. Runs are repeated multiple times and the low energy solutions are collected. Typically, annealing starts with qubits in superposition, but ends in or close to a ground state of the problem Hamiltonian.

Since the D-Wave machine relies upon the Ising/Hamiltonian model used in quantum annealing, the quadratic equation (1) above must be converted to the Ising/Hamiltonian formulation. This is readily accomplished using quadratic unconstrained binary optimization to express the four phi values (Figure 5) which are used instead of the four summed terms in red font in equation (1). Note that n1 and n2 in Figure 5 are analogous to x_1_ and x_2_ in Equation (1). The principle is exactly the same in the sense that we express the potential/weight functions in terms of the four permutations of the truth values of the flanking nodes.

**Figure 5:**
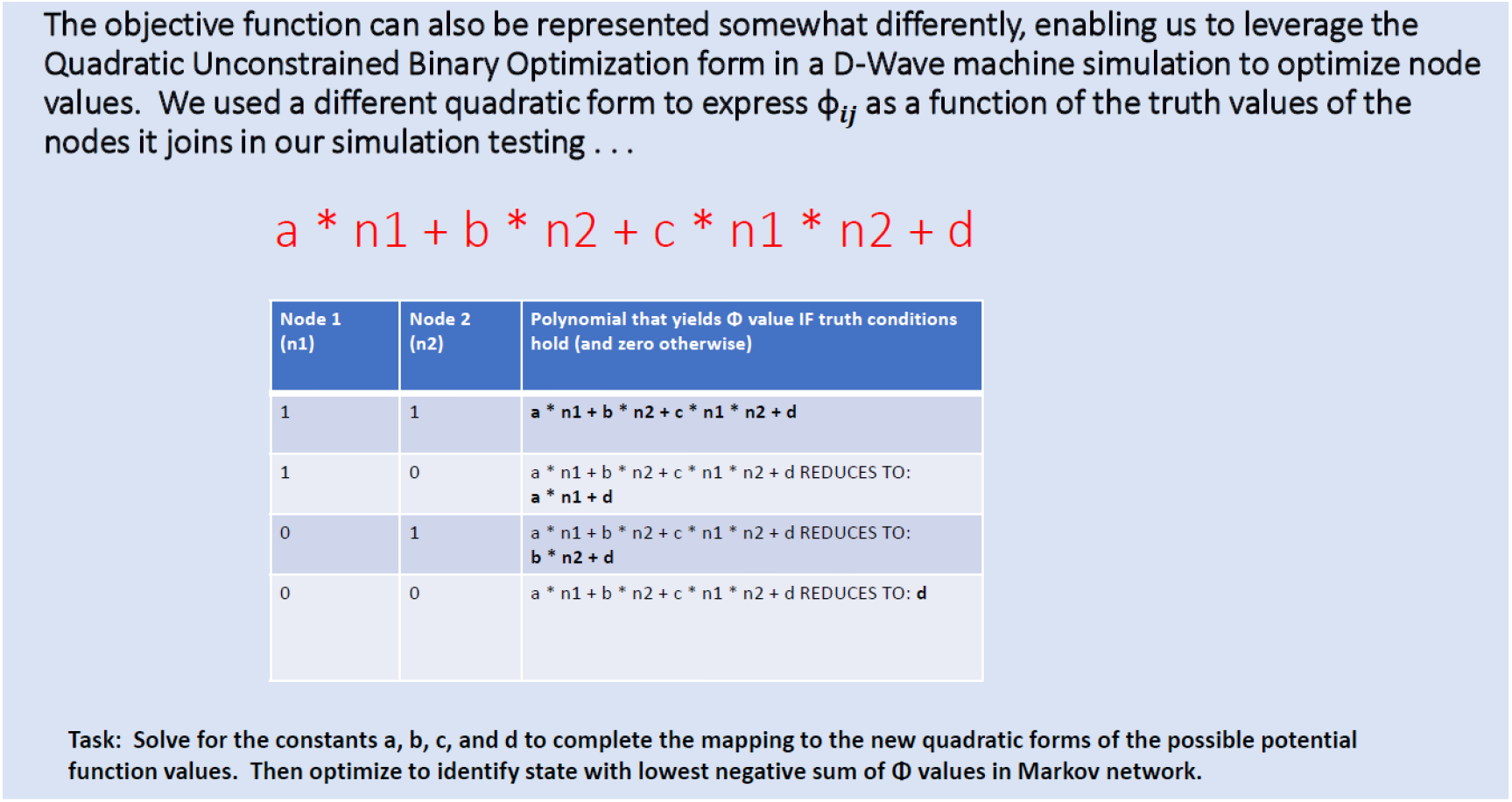
Expression of phi values using quadratic unconstrained binary optimization (QUBO)

## III. Proof-of Concept (POC) Objectives, Data Source, and Specific Methods

The objectives of our POC study were as follows:

1. Assess the accuracy of a relatively small-sized hybrid Markov Random Field-Bayesian graphical model in making multiple simultaneous clinical predictions:
  a. Development of bacteriuria in ICU (primary)
  b. Prediction of length of stay and other variables (secondary)
2. Evaluate correspondence between the predicted values in the most likely solution using classical inference (i.e., the most probable instantiation of all node values in the graph) versus quantum annealing (i.e., the lowest energy state)
3. Assess accuracy of quantum annealing-based predictions versus machine learning benchmarks

We constructed a graphical model based on MIMIC-III ICU database, which includes de-identified data for over 38,000 adult patients treated in intensive care units at Beth Israel Hospital in Boston between 2001 and 2012. This database includes ICD9 codes, past medical history, demographics, medications, laboratory results and other data.

Edges were placed between nodes that were linked probabilistically based on infectious diseases domain expertise. To accommodate data that were not available for certain variables, imputations were made using “Bayesian Islands” (see left side of Figure 6 below), which were parameterized using conditional probabilities from published literature and pre-solved (classically) prior to quantum annealing. The Markov Random Field (MRF) section of the graph (see right side of Figure 6) was order-reduced to pairwise connections and parameters (phi values informed by constants described in Figure 5) were learned using the pseudo-likelihood method based on data from a training set consisting of ∼95% of the evaluable data set (we also explored the use of a balanced training set). The remainder of the data were reserved as a validation/test set. In essence, once the Bayesian islands were solved, we were left with a pure MRF. The graph was embedded within the architecture of the D-Wave machine and, for each patient in the test set, the nodal truth value assignments (T/F) in the MRF portion of the graph were partially populated, withholding one or several (up to four) variables whose values were to be inferred using quantum annealing.

**Figure 6:**
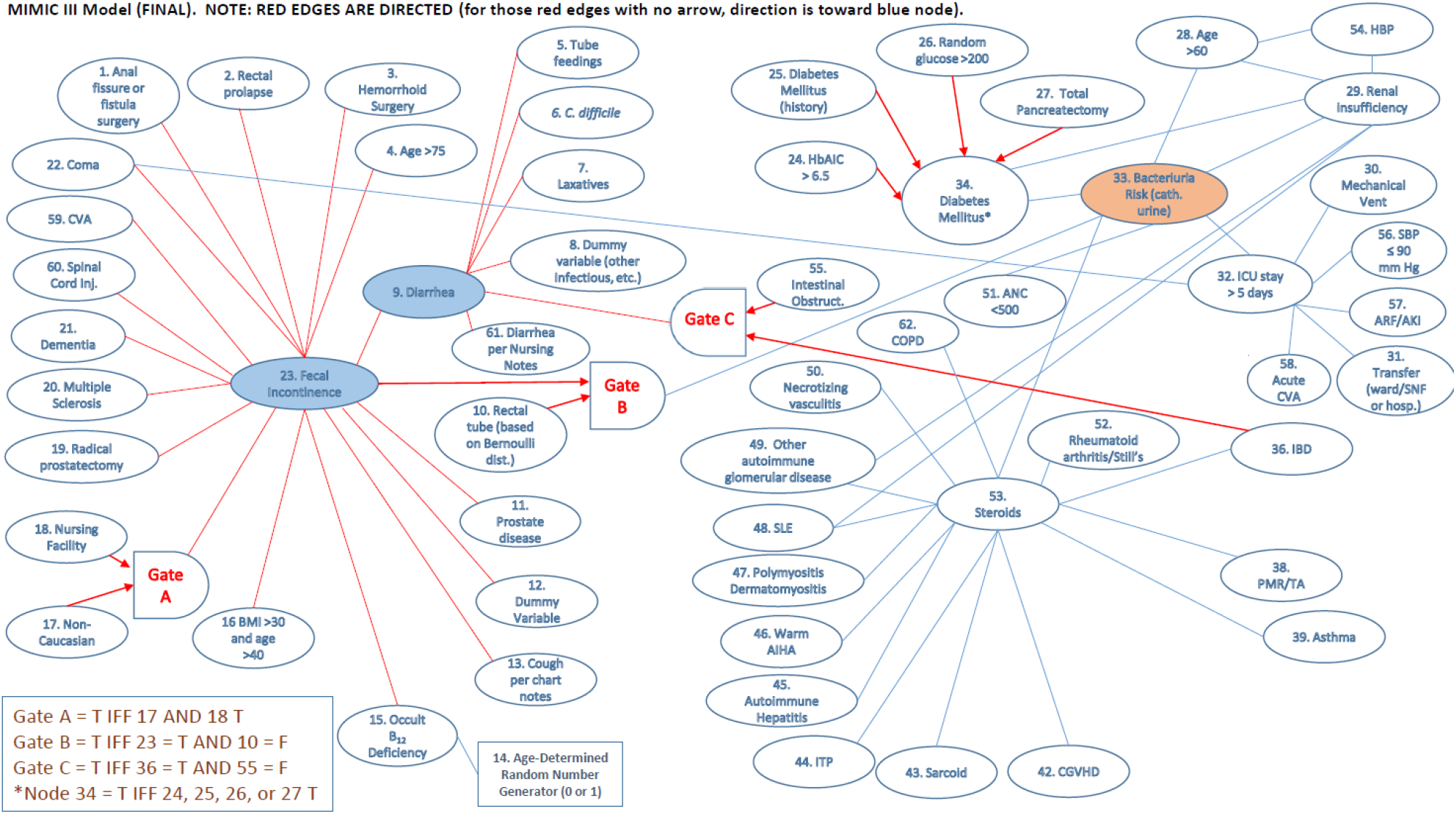
Topology of final model. Bayesian islands (pre-solved classically) indicated by directed (red) edges on left. Undirected edges of Markov Random Field portion of model in blue. The primary node to be predicted was node 33 (bacteriuria, or the development of a positive urine culture). During annealing runs, the truth values of up to four nodes were withheld for test patients and inferred results were compared with ground truth values for that patient in the database. Gates functioned as nodes with truth values imputed as described in the box in the lower left. Dummy variables were always set to true to subsume miscellaneous risk factors (e.g., for diarrhea in the ICU) not captured by other nodes. The age-determined truth value for Node 15 was based on a probability distribution, because, by definition, occult vitamin B_12_ deficiency was unknown [age <40: 1 = T in 2%, 40-59 (4%), 60-69 (5%), 70+ (6%)]. The “rectal tube AND fecal incontinence” node was populated using another random number generator (0=F vs. 1=T) based on a Bernoulli distribution informed by nursing note documentation indicating that 7.7% of patients with fecal incontinence had rectal tubes. Abbreviations: CVA (cerebrovascular accident), HBP (hypertension), SBP (systolic blood pressure), ARF/AKI (acute renal failure, acute kidney injury), ANC (absolute neutrophil count), COPD (chronic obstructive pulmonary disease), IBD (inflammatory bowel disease), SLE (systemic lupus erythematosus), PMR/TA (polymyalgia rheumatica/temporal arteritis), AHA (autoimmune hemolytic anemia), ITP (immune thrombocytopenia), CGVHD (chronic graft versus host disease). Several versions of the model were explored.

The objective function to be optimized incorporated phi values expressed in the “QUBO” form as outlined in Section II, and consisted of the negative value of the sum of the potential functions (phi values) for the pairwise cliques. The phi values in the objective function to be minimized were therefore based upon three quadratic edge coefficients and a constant (Figure 5) and the truth values of the adjoining nodes of the pair. Log transformation was accounted for in generating the values, enabling the use of a sum for the objective function. Quantum annealing runs were repeated up to 10,000 times to sample the solution space. Predictions informed by the lowest energy solution were compared with the known (i.e., withheld) values of these same variables in the MIMIC-III data set, enabling calculation of sensitivity, specificity and accuracy. We compared our results with those obtained using (classical) computation performed on a non-order reduced model (i.e., the “exact solution,” which was practical given the size of our graph), as well as several machine learning algorithms, including logistic regression and a feed-forward neural net.

## IV. Results of Proof-of-Concept

We compared the accuracy of predictions using quantum annealing versus those generated by classical exact inference in the subset of patients with confirmed Foley catheter placement for bladder drainage, a known risk factor for urinary tract infection. Data from a total of 18,977 patients was available, of whom 1000 patients were reserved for a validation/test set. Therefore, parameters (potential functions) for the MRF portion of the model were computed using data from approximately 17,977 patients.

Figure 7 below depicts accuracy, sensitivity, and specificity for classical inference (left panel) versus quantum annealing (right panel) when one, two, three, or four node values were withheld for test subjects.

**Figure 7:**
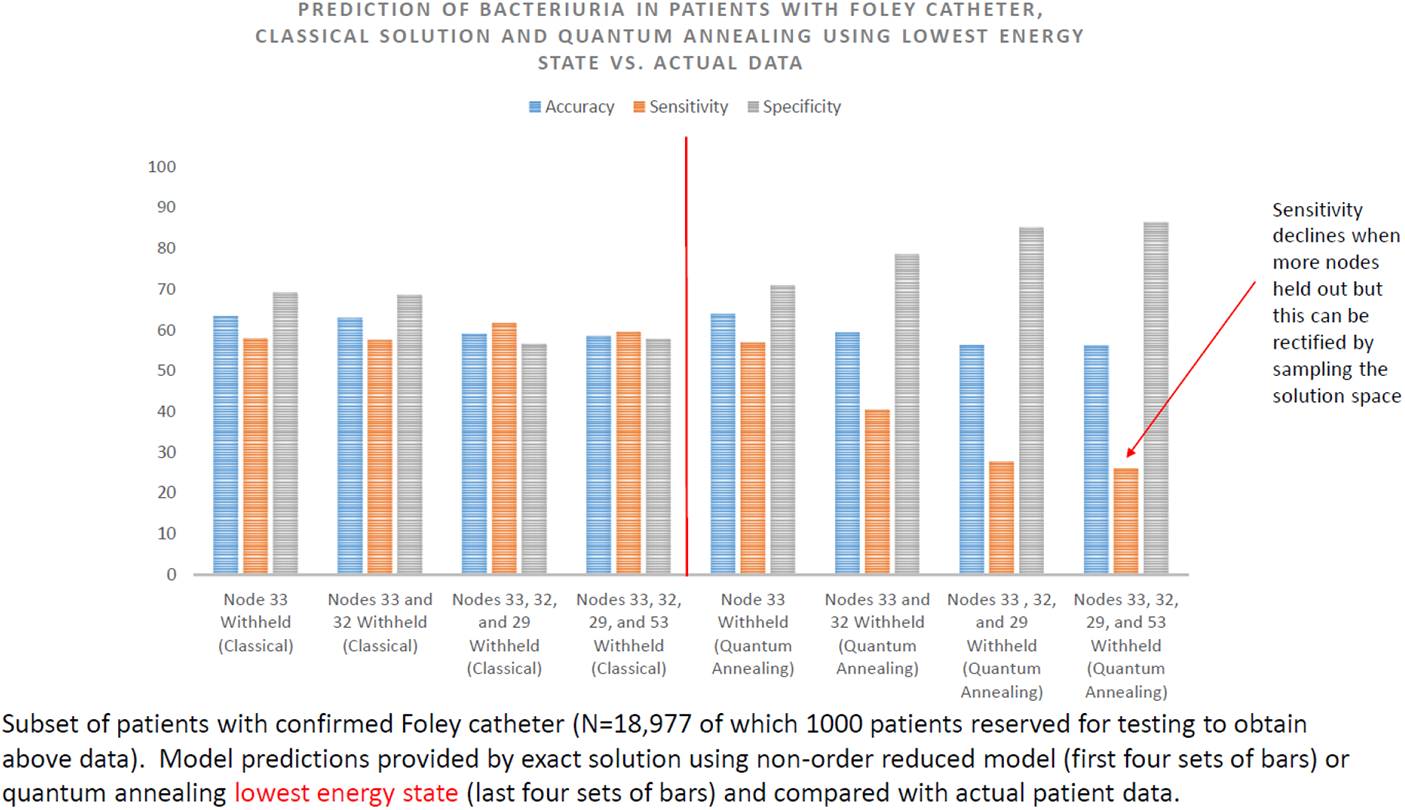
Summary of data for proof-of-concept study

In the primary analysis (prediction of bacteriuria alone), the results above demonstrate that the overall accuracy of quantum annealing (approximately 70%) is essentially identical to that of the gold standard of classical probabilistic inference (i.e., the exact solution based on the graphical model). Sensitivity was slightly higher than specificity. The moderate degree of predictive power was presumably related to the limitations of the data set, biological variability, and/or the topology of the graphical model, since these data show that performance was not compromised by application of a quantum annealing technique (at least in the primary analysis). Concordance of results (quantum annealing versus classical exact inference solution) was achieved in 99.7% of cases. Furthermore, performance of quantum annealing was comparable to that of machine learning benchmarks (feedforward neural net and logistic regression; data not shown).

Notably, as more nodes were withheld, quantum annealing sensitivity declined relative to that of classical probabilistic inference (that is, the exact solution). The reason for this behavior is not yet clear, although we found that this phenomenon was ameliorated when the solution space was more broadly sampled (evaluation of the 3 or 6 lowest energy solutions rather than a single presumed ground state solution; data not shown).

## V. Discussion and Ideas for Further Research

We have shown in a proof-of-concept study drawing upon data from approximately 19,000 patients that quantum annealing may serve as a predictive tool by enabling interrogation of a graphical model based on expert clinical knowledge and parameterized by population-based data. To our knowledge, this is the first time quantum annealing has been successfully deployed in this manner. In essence, optimization through annealing allowed us to solve for the most likely truth value of a withheld clinical variable (development of catheter-associated bacteriuria in the ICU). The modest predictive value of the model was not limited by the quantum technique when one variable was withheld, achieving, as it did, nearly complete concordance the classical inference solution. Preliminary data suggest, however, that in order to preserve sensitivity when multiple simultaneous predictions are made using this technique, it may be necessary to leverage additional data in the solution space rather than merely the lowest energy solution. This is an area for future research.

If this innovation successfully scales to larger models, one can easily envision broader applications, for example in a primary care capacity where practitioners must often care for complex patients with multiple co-morbidities who may have several issues that must be simultaneously addressed (i.e., more than one node for which it would be clinically useful to know the most likely truth value). In some cases – for example, the rank-ordering of personalized risk-benefit ratios for various treatment options – the approach may be modified, tapping into both classical and quantum computing in a series of orchestrated steps that involve repeated application of the model under various treatment scenarios followed by ranking (Fig 8).

**Figure 8:**
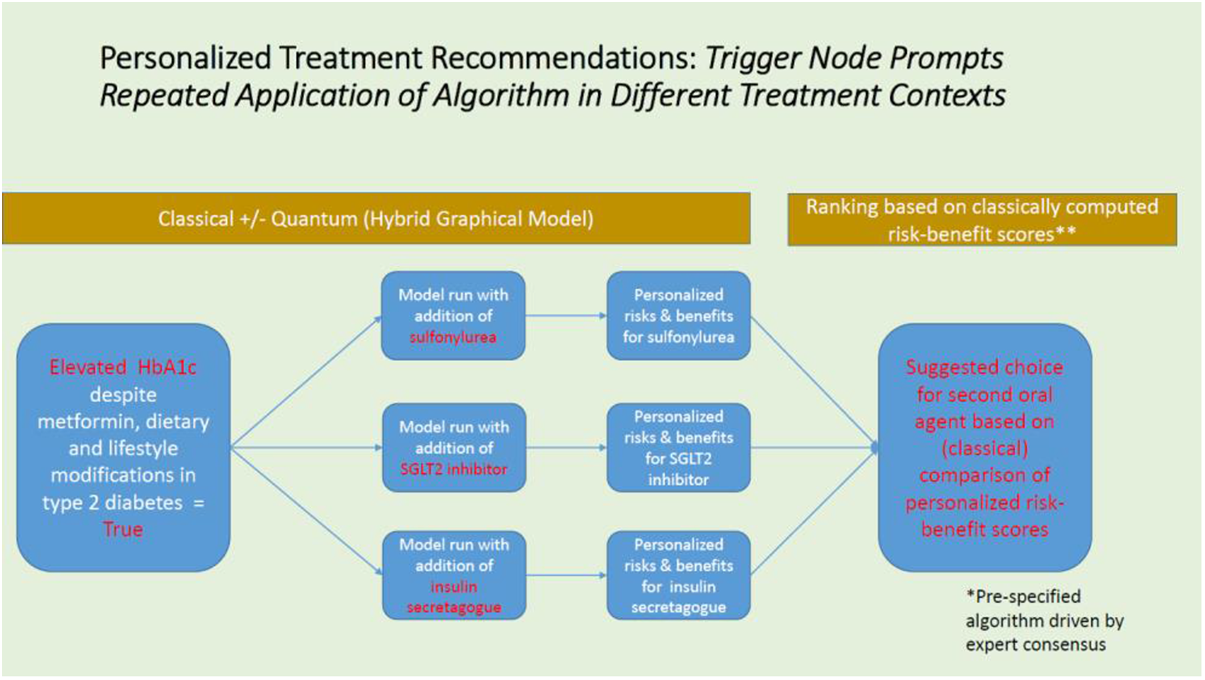
Hybrid approach

The D-wave service is cloud based, so the bulk of the “heavy lifting” is performed remotely. If appropriate interoperability is achieved with an electronic health record and other sources of data, the output of this system might funnel close-to-real time clinical decision support to busy primary care practitioners, struggling under a heavy caseload, on a hand-held device. In principle, the model should be updated and reparameterized on a regular basis with the accretion of more population-based data, especially given the pace of medical innovation. As currently envisioned, the system produces binary (dichotomous) outputs which represent the most likely truth values for unknown nodes based on the totality of the patient’s data and the existing parameterized model but more quantitative predictions may be possible through binning and adjunctive Bayesian methods. This, too, is an area rife for exploration.

## Data Availability

The MIMIC-III database that we used is publicly available.

## Author affiliations and conflict of interests

Neither author has a conflict of interest. David Sahner is the former Chief Scientific Officer and Chief Medical Officer of EigenMed, Inc., and now serves as a senior consultant to several biotechnology companies, providing support in clinical translation and machine learning. Richard Williams is the former Senior Vice President of Data Science at EigenMed, Inc.

## Acknowledgements

The authors would like to acknowledge Steve Reinhardt and Michael Booth (formerly of D-Wave) for their help in testing the algorithm, the late Peter Wittek of the Creative Destruction Lab affiliated with the University of Toronto for his encouragement, Eleanor Rieffel (NASA) for her helpful comments early in the program, and all of the former members of EigenMed, Inc., where this technology was refined, including Jon Mallon.

